## Supplementary Materials for "Noradrenergic deficits contribute to apathy in Parkinson’s disease through the precision of expected outcomes"

| | | |
|---|---|---|
| 1 | Mood and behaviour questionnaires | Page 1 |
| 2 | Relationship between prior weighting and apathy in young adults | Page 4 |
| 3 | Role of covariates in the drug × LC CNR interaction effect on prior weighting | Page 5 |
| 4 | Decomposing drug effects on prior weighting: plausible values analysis | Page 6 |
| 5 | Parameter recovery of prior weighting | Page 8 |
| 6 | Hierarchical Bayesian model specification | Page 11 |
| 7 | Comparing hierarchical Bayesian model variants | Page 12 |
| 8 | Posterior predictive checks of hierarchical Bayesian model | Page 13 |
| 9 | Supplementary references | Page 15 |

### MOOD AND BEHAVIOUR QUESTIONNAIRES

All participants completed the following set of self-report questionnaires that assessed mood and various behavioural symptoms: Apathy Scale (Starkstein et al., 1992), Barratt Impulsiveness Scale (Patton et al., 1995), Motivation and Energy Inventory (Fehnel et al., 2004), Hospital Anxiety and Depression Scale (Zigmond & Snaith, 1983), and Conners' Adult ADHD Rating Scale (Conners et al., 1999). Individuals with Parkinson's disease additionally completed the REM sleep behaviour disorder screening questionnaire (Stiasny-Kolster et al., 2007). Furthermore, relatives or friends of the Parkinson's disease patients completed informant-rated versions of the Conners' Adult ADHD Rating Scale and the Apathy Scale, as well as a general mood and behavioural symptom inventory (Cambridge Behavioural Inventory Revised; Wear et al., 2008).

**Table S1 | Descriptive statistics and group comparisons of questionnaires.**

| Measure | | PD | Controls | *BF* | *p* |
|---|---|---|---|---|---|
| Apathy Scale | Total Score (self-rated) | 12.68 (5.77) | 10.58 (5.09) | 0.58 | .212 |
| | Total Score (informant-rated) | 13.13 (5.59) | | | |
| BIS | Total Score | 56.45 (10.34) | 56.15 (9.67) | 0.3 | .924 |
| | Attention | 14.16 (4.3) | 14.23 (3.72) | 0.3 | .953 |
| | Motor | 20.08 (2.65) | 20.85 (3.38) | 0.39 | .398 |
| | Non-planning | 22.21 (5.4) | 21.08 (4.44) | 0.38 | .459 |
| HADS | Anxiety | 4.53 (3.2) | 4.31 (3.53) | 0.3 | .83 |
| | Depression | 3.95 (2.68) | 2.88 (2.76) | 0.58 | .202 |
| MEI | Total Score | 98.05 (21.3) | 108.96 (16.71) | 1.29 | .073 |
| | Mental | 44.11 (8.97) | 47.35 (8.09) | 0.57 | .22 |
| | Physical | 23.95 (6.95) | 29.35 (5.91) | 6.18 | .01 |
| | Social | 30 (7.34) | 32.27 (5.31) | 0.53 | .261 |
| CAARS (self-rated) | Inattention / Memory Problems | 5.42 (3.58) | 4.19 (3.06) | 0.55 | .235 |
| | Hyperactivity / Restlessness | 3.11 (2.71) | 2.77 (2.05) | 0.33 | .652 |
| | Impulsivity / Emotional Lability | 1.84 (1.5) | 2.77 (2.05) | 0.9 | .087 |
| | Problems with Self-Concept | 2.26 (2.33) | 3.88 (4.12) | 0.77 | .102 |
| | ADHD Index | 6.79 (4.26) | 8.38 (4.51) | 0.53 | .233 |

| | | |
|---|---|---|
| CAARS (observer-rated) | Inattention / Memory Problems | 4.24 (2.51) |
| | Hyperactivity / Restlessness | 1.58 (1.92) |
| | Impulsivity / Emotional Lability | 1.21 (1.23) |
| | Problems with Self-Concept | 2.32 (2.11) |
| | ADHD Index | 4.47 (3.42) |
| RBDSQ | | 4.58 (3.45) |
| CBI-R | Total Score | 15.13 (13.6) |
| | Abnormal Behaviour | 0.84 (1.12) |
| | Beliefs | 0.37 (1.21) |
| | Eating Habits | 0.95 (1.58) |
| | Everyday Skills | 1.16 (2.41) |
| | Memory and Orientation | 4.66 (3.9) |
| | Mood | 1.26 (2.1) |
| | Motivation | 2.26 (3.35) |
| | Stereotypic and Motor Behaviours | 0.79 (1.4) |
| | Self Care | 0.42 (0.84) |
| | Sleep | 2.42 (2.17) |

*Note*: Data are presented as mean (SD). Group comparisons were performed with independent samples t-tests. The stated *p*-values are uncorrected; none exceeded .05 after correction for multiple comparisons. Abbreviations: BF, Bayes Factor for the alternative hypothesis over the null hypothesis; BIS, Barratt Impulsiveness Scale; HADS, Hospital Anxiety and Depression Scale; MEI, Motivation and Energy Inventory; CAARS, Conners' Adult ADHD Rating Scale; RBDSQ, REM sleep Behaviour Disorder Screening Questionnaire; CBI-R, Cambridge Behavioural Inventory – Revised.

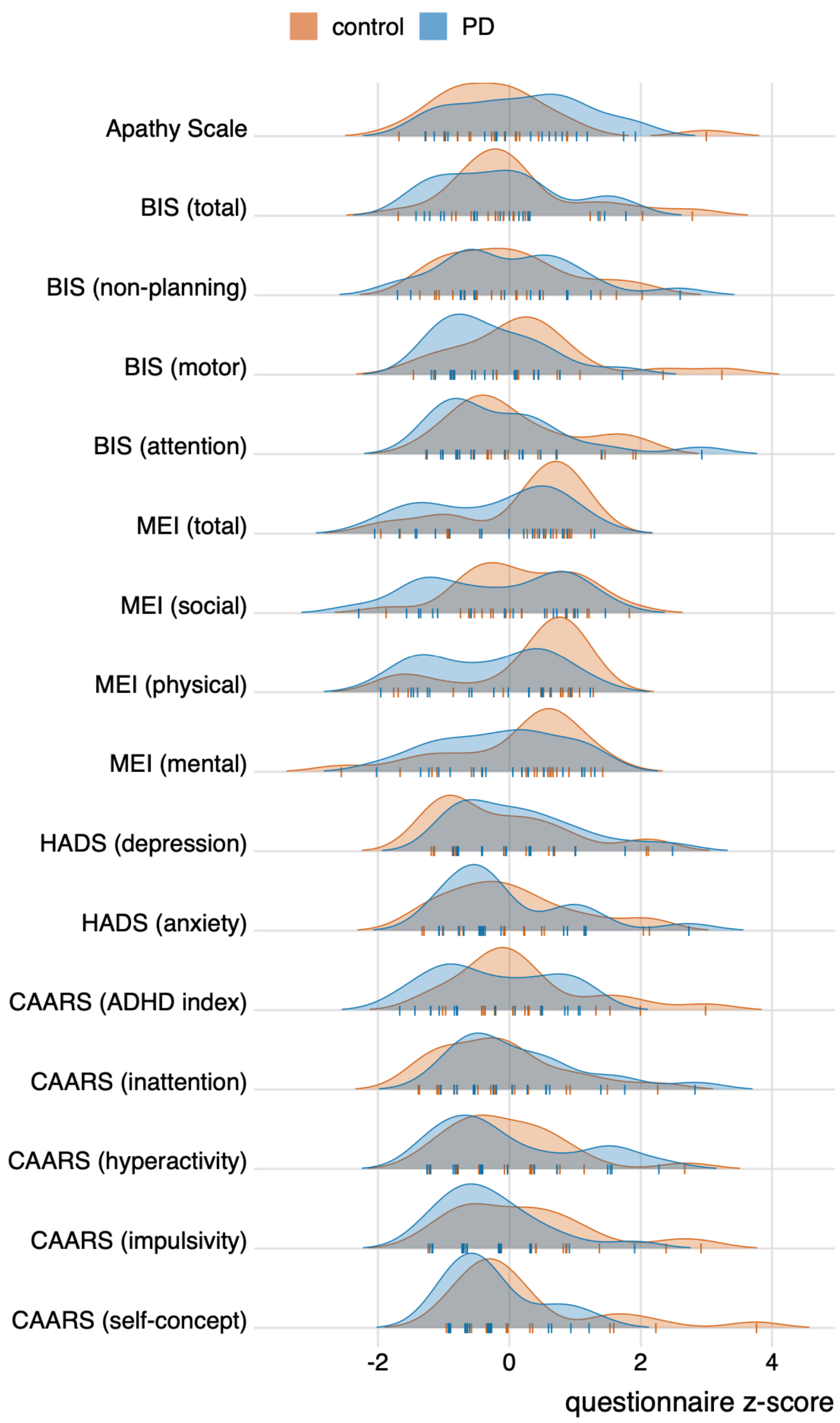

**Figure S1 | Density plots of questionnaire scores for participants with Parkinson's disease (blue) and controls (orange).** The questionnaire scores were z-scored to bring the different questionnaires onto a common scale (note that this transformation does not affect group comparisons for a given questionnaire outcome). Tick marks reflect individual data points. Abbreviations: BIS, Barratt Impulsiveness Scale; MEI, Motivation and Energy Inventory; HADS, Hospital Anxiety and Depression Scale; CAARS, Conners' Adult ADHD Rating Scale.

### RELATIONSHIP BETWEEN PRIOR WEIGHTING AND APATHY IN YOUNG ADULTS

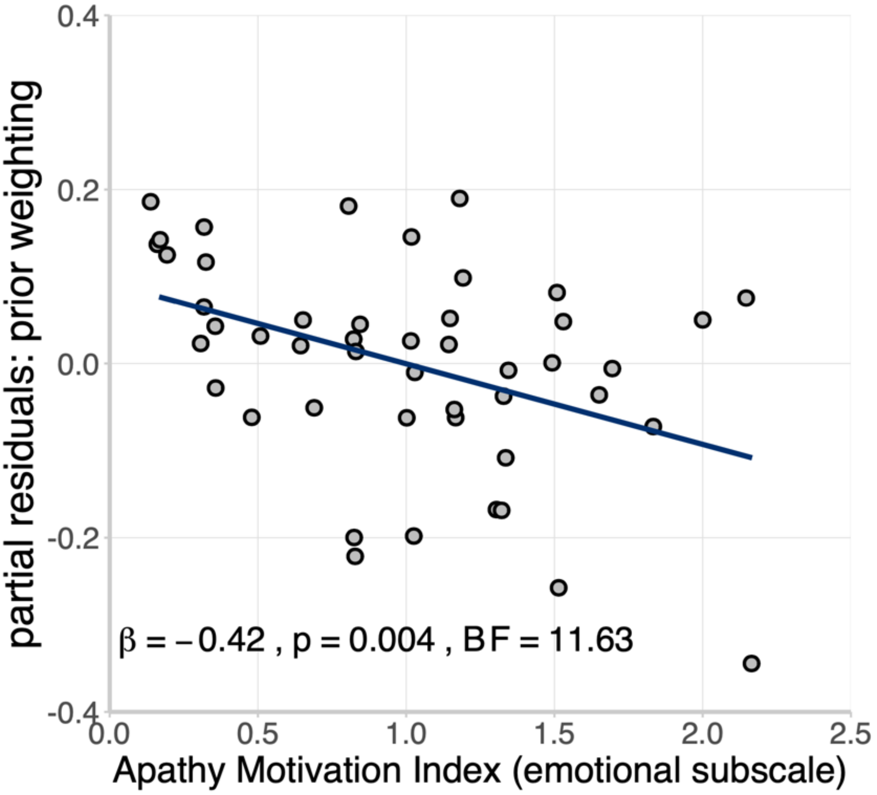

**Figure S2 | Prior weighting and trait apathy in young adults.** Data from Hezemans et al. (2020), demonstrating the relationship between trait apathy (measured using the Apathy Motivation Index; Ang et al., 2017) and prior weighting, adjusted for task performance variability (i.e., partial residuals). Note that observations with identical questionnaire scores were horizontally jittered to avoid overlaid dots. Full statistics for the regression coefficient of apathy: $\beta = -0.42$, SE = 0.14, $t_{(44)} = -3.02$, $p = .004$; BF = 11.63.

### ROLE OF COVARIATES IN THE DRUG × LC CNR INTERACTION ON PRIOR WEIGHTING

**Table S2 | Backward elimination of fixed effects in the linear mixed effects model predicting prior weighting.**

| | Model selection step | | | | | |
|---|---|---|---|---|---|---|
| **Predictors** | 1 | 2 | 3 | 4 | 5 | 6 |
| Drug | $-2.02 \times 10^{-3}$ | $-2.02 \times 10^{-3}$ | $-2.06 \times 10^{-3}$ | $-2.06 \times 10^{-3}$ | $-2.06 \times 10^{-3}$ | $-2.06 \times 10^{-3}$ |
| LC CNR | -0.02 | -0.02 | -0.02 | -0.06 | -0.08 | -0.13 |
| Drug × LC CNR | -0.37* | -0.37* | -0.38* | -0.38* | -0.38* | -0.38* |
| ICV | 0.37 | 0.37 | 0.37 | 0.37 | 0.35 | 0.37* |
| Age | 0.22 | 0.23 | 0.23 | 0.19 | 0.18 | - |
| Ato plasma | -0.13 | -0.13 | -0.13 | -0.11 | - | - |
| LEDD | 0.13 | 0.14 | 0.14 | - | - | - |
| Session | 0.07 | 0.07 | - | - | - | - |
| UPDRS III | 0.01 | - | - | - | - | - |
| **Information Criteria** | | | | | | |
| AIC | 100.59 | 98.59 | 96.89 | 95.77 | 94.38 | 93.69 |
| Δ AIC | 6.89 | 4.90 | 3.20 | 2.07 | 0.68 | 0 |
| BIC | 120.43 | 116.91 | 113.68 | 111.03 | 108.12 | 105.91 |
| Δ BIC | 14.52 | 11.00 | 7.78 | 5.12 | 2.21 | 0 |

*Note.* Values for predictors are standardised regression coefficients (ß). *$p < .05$. Drug, atomoxetine vs. placebo condition; LC CNR, Locus Coeruleus Contrast to Noise Ratio; ICV, total intracranial volume; Ato plasma, atomoxetine plasma concentration; LEDD, Levodopa Equivalent Daily Dose; Session, first vs. second session; UPDRS III, Unified Parkinson's Disease Rating Scale, motor examination; AIC, Akaike Information Criterion; BIC, Bayesian Information Criterion; Δ AIC / BIC, difference in AIC / BIC with respect to the lowest AIC / BIC value. All models included a fixed effect of the interquartile range of performance error as a covariate of no interest, and a random effect of participants on the intercept. Total intracranial volume was estimated from the T1-weighted MP2RAGE images using the `mri_segstats –etiv-only` procedure in FreeSurfer v6.0.0 (Malone et al., 2015).

**Table S3 | Bayes Factors for the inclusion of fixed effects in the linear mixed effects model predicting prior weighting.**

| **Predictors** | P(incl) | P(incl\|data) | P(excl\|data) | $BF_{inclusion}$ |
|---|---|---|---|---|
| Drug | 0.40 | 0.13 | 0.40 | 0.32 |
| LC CNR | 0.40 | 0.19 | 0.33 | 0.59 |
| Drug × LC CNR | 0.20 | 0.48 | 0.05 | 10.06 |
| ICV | 0.50 | 0.58 | 0.42 | 1.40 |
| Age | 0.50 | 0.42 | 0.58 | 0.72 |
| Ato plasma | 0.50 | 0.38 | 0.62 | 0.62 |
| LEDD | 0.50 | 0.38 | 0.62 | 0.61 |
| Session | 0.50 | 0.30 | 0.70 | 0.43 |
| UPDRS III | 0.50 | 0.40 | 0.60 | 0.67 |

*Note.* P(incl): prior inclusion probability, i.e. the summed prior probability of models that include the predictor. A priori, all possible restrictions of the full model were deemed to be equally likely (i.e., a uniform prior was assigned to the model space). Thus, P(incl) reflects the proportion of alternative models that included the predictor. P(incl|data): posterior inclusion probability, i.e. the summed posterior probability of models that include the predictor. P(excl|data): posterior exclusion probability, i.e. the summed posterior probability of models that exclude the predictor. $BF_{inclusion}$: Inclusion Bayes Factor, i.e. the change from prior to posterior inclusion odds. This indicates how much more likely the data are under models that include the predictor, compared to models that exclude the predictor (Hinne et al., 2020). This analysis was performed using "matched" models, which means that (i) models were not permitted to include an interaction effect without its constituent main effects, and (ii) inclusion probabilities for an interaction effect were based only on the subset of models that contained (at least) the constituent main effects of the interaction (Mathôt, 2017). All models included a fixed effect of the interquartile range of performance error as a covariate of no interest, and a random effect of participants on the intercept.

### Decomposing drug effects on prior weighting: plausible values analysis

In the main manuscript, we examined whether the effect of atomoxetine on prior weighting was differentially associated with the effects of atomoxetine on prior precision and sensory evidence precision. The drug effects on prior and sensory evidence precision were estimated using hierarchical Bayesian estimation of a model of perceptual inference (Figure S4). Hierarchical techniques fit the data from all participants simultaneously, while explicitly accounting for individual differences. Compared to fitting each participant separately, hierarchical modelling produces participant-level parameter estimates that are – on average – closer to the true parameter values, due to shrinkage towards the mean (Farrell & Lewandowsky, 2018, pp. 203–238). Although this shrinkage is advantageous in the context of parameter estimation, it can be problematic for post-hoc statistical testing. Specifically, because participant-level parameter estimates from hierarchical models are shrunk towards each other, statistical analyses that pool across these parameter estimates will underestimate the pooled standard deviation, leading to inflated effect sizes and test statistics (Boehm et al., 2018).

To address this issue, we performed a plausible values analysis that accounts for the uncertainty of the participant-level parameter estimates from the hierarchical model (Ly et al., 2017). This approach repeats the statistical test of interest for each of the Markov Chain Monte Carlo (MCMC) samples from the joint posterior distribution of the hierarchical model. In other words, the statistical test is repeated for each set of plausible values of the parameters, rather than relying on a single summary of the parameters (e.g., posterior medians). This yields a full posterior distribution of the test statistic, which explicitly represents the estimation uncertainty.

We obtained the distributions of plausible correlations between the drug effect on prior weighting and the estimated drug effects on prior precision (Figure S3A) and sensory evidence precision (Figure S3B). There was a strongly negative correlation between the drug effect on prior weighting and the drug effect on the standard deviation of the prior, with the distribution of plausible correlations reliably shifted below zero (median = -0.55, 95% QI: [-0.76, -0.27], $P(r < 0) = 99.98\%$). In contrast, the distribution of plausible correlations between the drug effect on prior weighting and the drug effect on the standard deviation of sensory evidence was approximately centred on zero (median = -0.04, 95% QI: [-0.37, 0.34], $P(r < 0) = 57.73\%$). To examine whether these correlations were different from each other, we calculated the difference between the plausible correlation estimates for each MCMC sample. The distribution of the difference between correlations was clearly shifted below zero (Figure S3C; median = -0.51, 95% QI: [-0.93, -0.09], $P(\Delta r < 0) =$

99.18%), indicating that the correlation with the drug effect on prior precision was significantly stronger (i.e., more negative) than the correlation with the drug effect on sensory evidence precision.

Taken together, the results from this supplementary analysis are consistent with the results presented in the main manuscript, suggesting that the atomoxetine-induced change in prior weighting was primarily explained by changes in prior precision, and not by changes in sensory evidence precision.

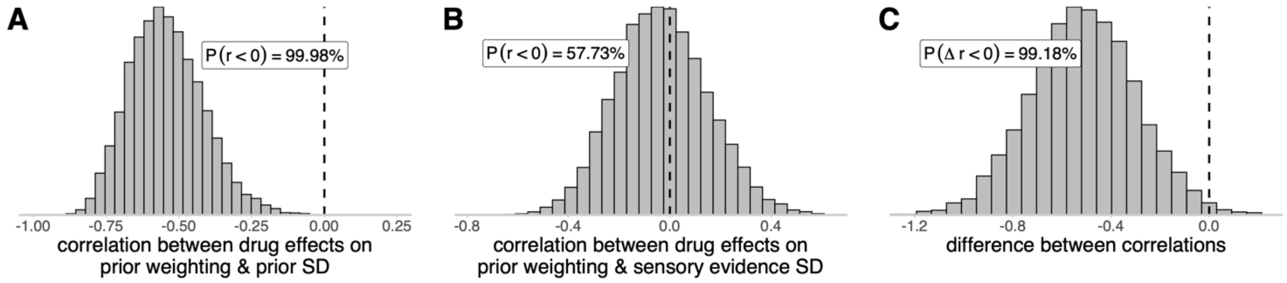

**Figure S3 | Decomposing the noradrenergic effects on prior weighting using a plausible values analysis approach.** (A-B) Distributions of plausible correlations between the drug effect on prior weighting and the estimated drug effect on the standard deviation of the prior (A) or sensory evidence (B). (C) Distribution of the difference between the plausible correlations.

### PARAMETER RECOVERY OF PRIOR WEIGHTING

We performed a parameter recovery analysis to examine the identifiability of prior weighting, as estimated by a linear mixed effects model of estimation error against performance error. To this end, we first simulated 100 datasets of performance errors, closely following the design of the visuomotor task. Each simulated dataset consisted of four blocks of 30 trials, with one block for each condition of the effort and reward manipulations. For each simulated dataset $i$ and condition $k$, the vector of 30 simulated performance errors $\boldsymbol{x}_{\text{sim}}^{(i,k)}$ was drawn from a truncated normal distribution:

$$\boldsymbol{x}_{\text{sim}}^{(i,k)} \sim \mathcal{N}_{(x_{\min}, x_{\max})}\left(\mu^{(k)}, \sigma^{(k)}\right) \tag{S1}$$

where $\mu^{(k)}$ and $\sigma^{(k)}$ represent the observed group-level mean and standard deviation of performance error for condition $k$, and the truncation points $x_{\min}$ and $x_{\max}$ represent the bounds on performance error, given the target position for condition $k$ and the limits of the laptop screen.

We then assigned the standard deviations of the prior and sensory evidence distributions, $\sigma_{\text{prior}}^{(i)}$ and $\sigma_{\text{evidence}}^{(i)}$, which jointly constitute the prior weighting term $w_{\text{prior}}^{(i)}$ and the posterior standard deviation $\sigma_{\hat{x}}^{(i)}$. Given that the standard deviations of the prior and sensory evidence are strongly correlated with the variance of performance errors (Hezemans et al., 2020; Wolpe et al., 2014), they were drawn from truncated normal distributions whose mean and standard deviation depended on the standard deviation of the simulated performance errors, $\sigma_{\text{sim}}^{(i)}$:

$$\begin{gathered}
\sigma_{\text{prior}}^{(i)} \sim \mathcal{N}_{+}\left(\sigma_{\text{sim}}^{(i)}, 0.5 \cdot \sigma_{\text{sim}}^{(i)}\right) \\
\sigma_{\text{evidence}}^{(i)} \sim \mathcal{N}_{\left(0.5 \cdot \sigma_{\text{sim}}^{(i)}, 1.5 \cdot \sigma_{\text{sim}}^{(i)}\right)}\left(\sigma_{\text{sim}}^{(i)}, 0.3 \cdot \sigma_{\text{sim}}^{(i)}\right) \\
w_{\text{prior}}^{(i)} = \frac{\left[\sigma_{\text{evidence}}^{(i)}\right]^2}{\left[\sigma_{\text{evidence}}^{(i)}\right]^2 + \left[\sigma_{\text{prior}}^{(i)}\right]^2} \\
\sigma_{\hat{x}}^{(i)} = \frac{\left[\sigma_{\text{evidence}}^{(i)}\right]^2 \cdot \left[\sigma_{\text{prior}}^{(i)}\right]^2}{\left[\sigma_{\text{evidence}}^{(i)}\right]^2 + \left[\sigma_{\text{prior}}^{(i)}\right]^2}
\end{gathered} \tag{S2}$$

Note that we constrained the standard deviation of sensory evidence to be within a relatively close range of the standard deviation of the simulated performance errors, $\left[0.5 \cdot \sigma_{\text{sim}}^{(i)}, 1.5 \cdot \sigma_{\text{sim}}^{(i)}\right]$, since we expected individual differences in prior weighting to be primarily driven by variation in the standard deviation of the prior (Hezemans et al., 2020).

For each simulated dataset $i$, we pseudorandomly selected 10 trials within each block $k$ as estimation trials, thus yielding 40 estimation trials. For a given estimation trial $n$, the optimal estimate of the final ball position is a precision-weighted combination of the prior (centred on the target position) and the likelihood (centred on the simulated true final ball position):

$$\hat{x}^{(i,n)} = w_{\text{prior}}^{(i)} \cdot x_{\text{target}}^{(i,k)} + \left(1 - w_{\text{prior}}^{(i)}\right) \cdot x_{\text{ball}}^{(i,n)} \tag{S3}$$

Considering the posterior standard deviation of this optimal estimate, $\sigma_{\hat{x}}^{(i)}$, we simulated the estimated final ball position by drawing from the trial-wise posterior distribution:

$$x_{\text{estimate}}^{(i,n)} \sim \mathcal{N}\left(\hat{x}^{(i,n)}, \sigma_{\hat{x}}^{(i)}\right) \tag{S4}$$

In line with our task design, we pseudorandomly determined the position of an estimation grid, consisting of a set of 12 evenly spaced response options $R$, for each estimation trial $n$. The simulated response $y$ for estimation trial $n$ is the response option $r$ that minimises the absolute distance from the simulated estimate of the final ball position:

$$y^{(i,n)} = \underset{r \in R}{\operatorname{argmin}} \left| r - x_{\text{estimate}}^{(i,n)} \right| \tag{S5}$$

The estimation errors are then given as the difference between the simulated response and the response option centred on the true final ball position. The z-scored estimation errors were regressed against the z-scored performance errors, and the negative of the regression coefficient was taken as the estimate of prior weighting. To examine the uncertainty of parameter recovery, we repeated the simulation of the estimated final ball position (Equation S4) 2000 times. Thus, for each data-generating setting of prior weighting $w_{\text{prior}}^{(i)}$ and the corresponding vector of $N$ simulated estimation trials, we obtained a $2000 \times N$ matrix of plausible estimates of the final ball positions. We performed the regression analysis of estimation error against performance error separately for each of the 2000 repetitions, yielding a distribution of plausible estimates of prior weighting for each of the 100 data-generating prior weighting values.

Parameter recovery performance is illustrated below in Figure S4. The data-generating and median estimated prior-weighting values were very strongly correlated ($r_{(98)} = 0.94$, $t = 27.37$, $p < .001$; $BF = 3.54 \times 10^{42}$), indicating good parameter recovery. There was a small positive bias, such that the estimates of prior weighting tended to be slightly higher than the data-generating values (mean bias = 0.04, SD = 0.08).

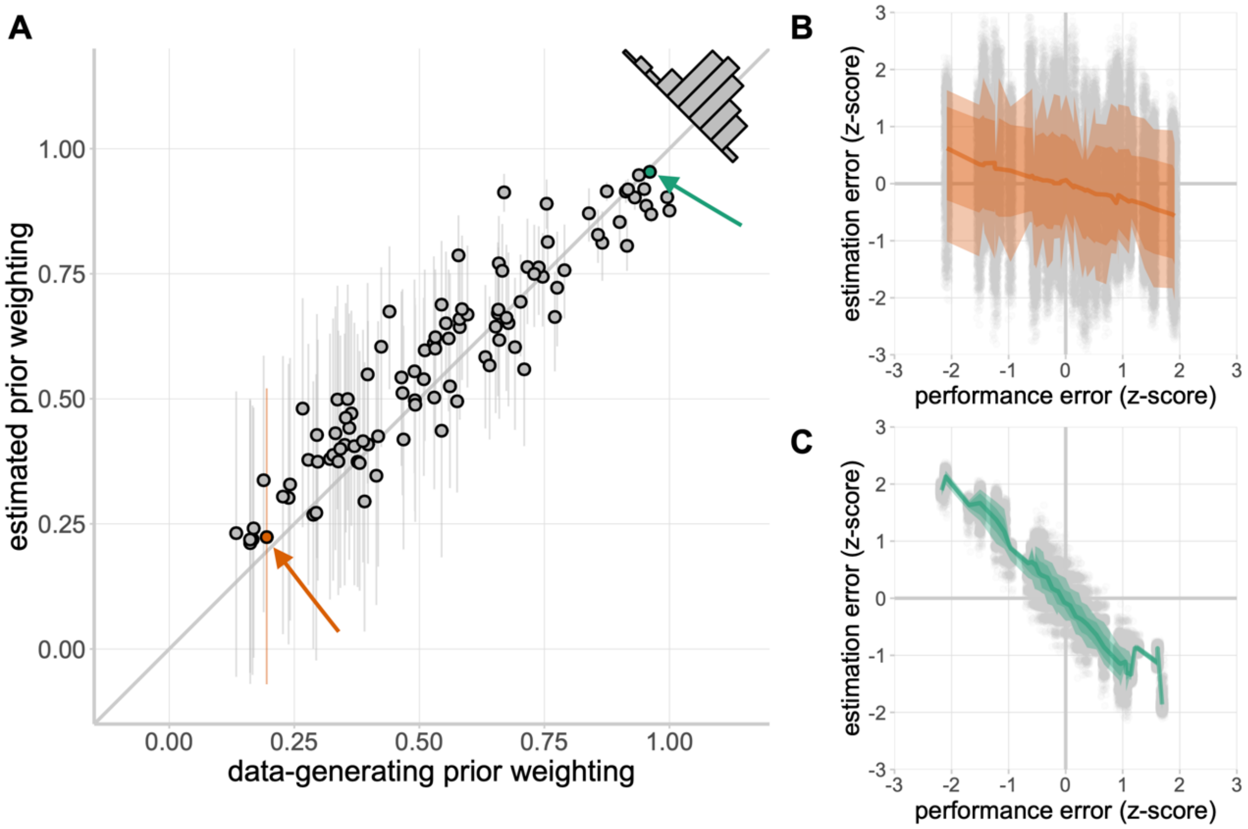

**Figure S4 | Parameter recovery of prior weighting.** (A) Data-generating prior weighting plotted against the estimated (i.e., recovered) prior weighting. The diagonal (identity) line represents perfect parameter recovery. For a given data-generating prior weighting value, the dot represents the median of the prior weighting estimates across 2000 simulations of estimation errors; the vertical error bars represent the 95% quantile intervals of the prior weighting estimates. The inset histogram illustrates the distribution of the difference between the median estimated prior weighting and data-generating prior weighting. (B, C) Examples of data simulations for a relatively low data-generating value of prior weighting (B; orange dot and error bar in A) and a relatively high data-generating value of prior weighting (C; green dot in A).

### HIERARCHICAL BAYESIAN MODEL SPECIFICATION

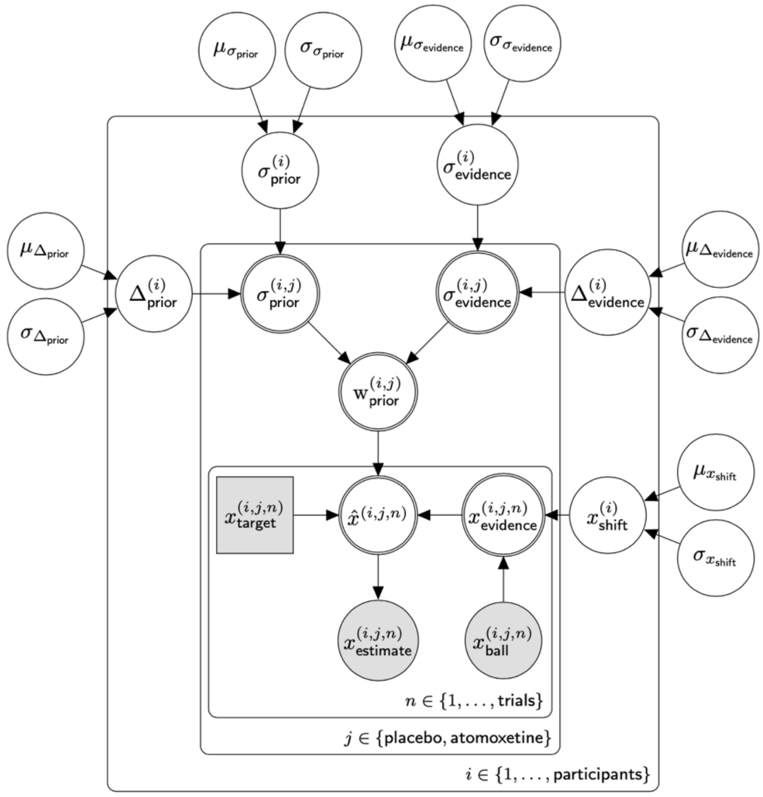

group-level prior SD:

$$\mu_{\sigma_{\text{prior}}} \sim \mathcal{N}(100, 10) \qquad \mu_{\sigma_{\text{prior}}} > 0$$
$$\sigma_{\sigma_{\text{prior}}} \sim \text{Cauchy}(0, 5) \qquad \sigma_{\sigma_{\text{prior}}} > 0$$

group-level drug effect on prior SD:

$$\mu_{\Delta_{\text{prior}}} \sim \mathcal{N}(0, 10)$$
$$\sigma_{\Delta_{\text{prior}}} \sim \text{Cauchy}(0, 5) \qquad \sigma_{\Delta_{\text{prior}}} > 0$$

group-level sensory evidence SD:

$$\mu_{\sigma_{\text{evidence}}} \sim \mathcal{N}(100, 10) \qquad \mu_{\sigma_{\text{evidence}}} > 0$$
$$\sigma_{\sigma_{\text{evidence}}} \sim \text{Cauchy}(0, 5) \qquad \sigma_{\sigma_{\text{evidence}}} > 0$$

group-level drug effect on sensory evidence SD:

$$\mu_{\Delta_{\text{evidence}}} \sim \mathcal{N}(0, 10)$$
$$\sigma_{\Delta_{\text{evidence}}} \sim \text{Cauchy}(0, 5) \qquad \sigma_{\Delta_{\text{evidence}}} > 0$$

group-level sensory evidence shift:

$$\mu_{x_{\text{shift}}} \sim \mathcal{N}(0, 10)$$
$$\sigma_{x_{\text{shift}}} \sim \text{Cauchy}(0, 5) \qquad \sigma_{x_{\text{shift}}} > 0$$

---

participant-level prior SD:

$$\sigma^{(i)}_{\text{prior}} \sim \mathcal{N}\left(\mu_{\sigma_{\text{prior}}}, \sigma_{\sigma_{\text{prior}}}\right) \qquad \sigma^{(i)}_{\text{prior}} > 0$$

participant-level drug effect on prior SD:

$$\Delta^{(i)}_{\text{prior}} \sim \mathcal{N}\left(\mu_{\Delta_{\text{prior}}}, \sigma_{\Delta_{\text{prior}}}\right)$$

participant-level sensory evidence SD:

$$\sigma^{(i)}_{\text{evidence}} \sim \mathcal{N}\left(\mu_{\sigma_{\text{evidence}}}, \sigma_{\sigma_{\text{evidence}}}\right) \qquad \sigma^{(i)}_{\text{evidence}} > 0$$

participant-level drug effect on sensory evidence SD:

$$\Delta^{(i)}_{\text{evidence}} \sim \mathcal{N}\left(\mu_{\Delta_{\text{evidence}}}, \sigma_{\Delta_{\text{evidence}}}\right)$$

participant-level sensory evidence shift:

$$x^{(i)}_{\text{shift}} \sim \mathcal{N}\left(\mu_{x_{\text{shift}}}, \sigma_{x_{\text{shift}}}\right)$$

---

session-level prior SD:

$$\sigma^{(i,j)}_{\text{prior}} \leftarrow \begin{cases} \sigma^{(i)}_{\text{prior}}, & \text{if } j = \text{placebo} \\ \sigma^{(i)}_{\text{prior}} + \Delta^{(i)}_{\text{prior}}, & \text{if } j = \text{atomoxetine} \end{cases}$$

session-level sensory evidence SD:

$$\sigma^{(i,j)}_{\text{evidence}} \leftarrow \begin{cases} \sigma^{(i)}_{\text{evidence}}, & \text{if } j = \text{placebo} \\ \sigma^{(i)}_{\text{evidence}} + \Delta^{(i)}_{\text{evidence}}, & \text{if } j = \text{atomoxetine} \end{cases}$$

session-level prior weighting:

$$\text{w}^{(i,j)}_{\text{prior}} \leftarrow \frac{\left[\sigma^{(i,j)}_{\text{evidence}}\right]^2}{\left[\sigma^{(i,j)}_{\text{evidence}}\right]^2 + \left[\sigma^{(i,j)}_{\text{prior}}\right]^2}$$

session-level posterior SD:

$$\sigma^{(i,j)}_{\hat{x}} \leftarrow \sqrt{\frac{\left[\sigma^{(i,j)}_{\text{evidence}}\right]^2 \cdot \left[\sigma^{(i,j)}_{\text{prior}}\right]^2}{\left[\sigma^{(i,j)}_{\text{evidence}}\right]^2 + \left[\sigma^{(i,j)}_{\text{prior}}\right]^2}}$$

---

trial-level sensory evidence mean:

$$x^{(i,j,n)}_{\text{evidence}} \leftarrow x^{(i,j,n)}_{\text{ball}} + x^{(i)}_{\text{shift}}$$

trial-level posterior mean:

$$\hat{x}^{(i,j,n)} \leftarrow \text{w}^{(i,j)}_{\text{prior}} \cdot x^{(i,j,n)}_{\text{target}} + \left[1 - \text{w}^{(i,j)}_{\text{prior}}\right] \cdot x^{(i,j,n)}_{\text{evidence}}$$

trial-level estimate of performance:

$$x^{(i,j,n)}_{\text{estimate}} \sim \mathcal{N}\left(\hat{x}^{(i,j,n)}, \sigma^{(i,j)}_{\hat{x}}\right)$$

**Figure S5 | Plate notation and sampling statements for the hierarchical Bayesian model.** Participant-level parameters were sampled from latent group-level distributions. We allowed for atomoxetine-induced changes in the standard deviations of the prior and sensory evidence distributions. Session-level parameters were then used to obtain trial-level posterior distributions of the final ball position. The observed estimates of performance were modelled as samples from the trial-level posterior distributions. The model is represented in plate notation: shaded nodes represent observed data whereas white nodes represent latent variables; rectangular nodes represent discrete or fixed variables whereas circular nodes represent continuous variables; and double-bordered white nodes represent deterministic variables whereas single-bordered white nodes represent stochastic variables.

### COMPARING HIERARCHICAL BAYESIAN MODEL VARIANTS

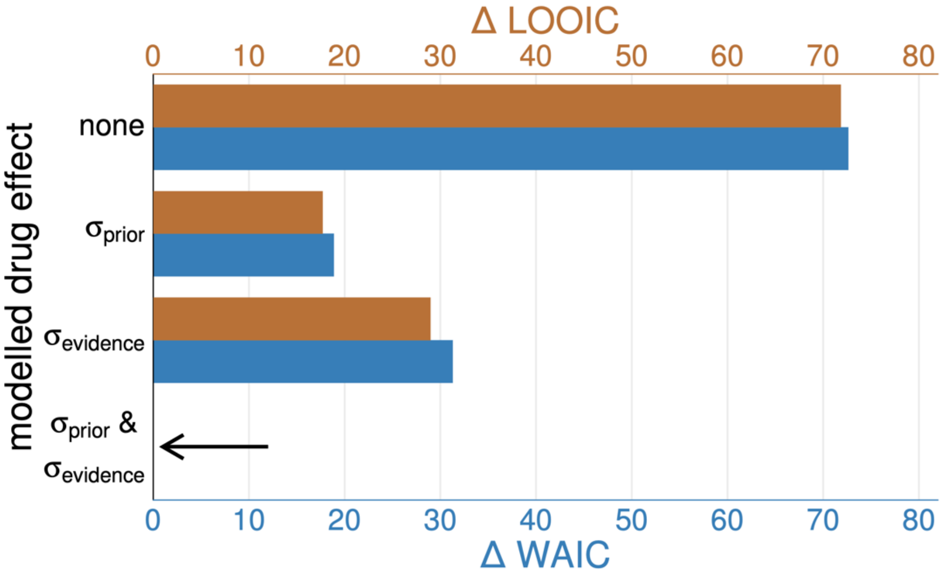

**Figure S6 | Information criteria for variants of the hierarchical Bayesian model.** In addition to the model presented in Figure S5, we fit three variants of the model with restrictions on the number of drug-induced change parameters. For each model variant, we computed the leave-one-out information criterion (LOOIC) and the widely applicable information criterion (WAIC) as estimates of the model's expected predictive accuracy (Vehtari et al., 2017). For both measures, the 'full' model which included drug-induced change parameters for both the prior and sensory evidence standard deviations was strongly preferred over the three more restricted model variants.

### POSTERIOR PREDICTIVE CHECKS OF HIERARCHICAL BAYESIAN MODEL

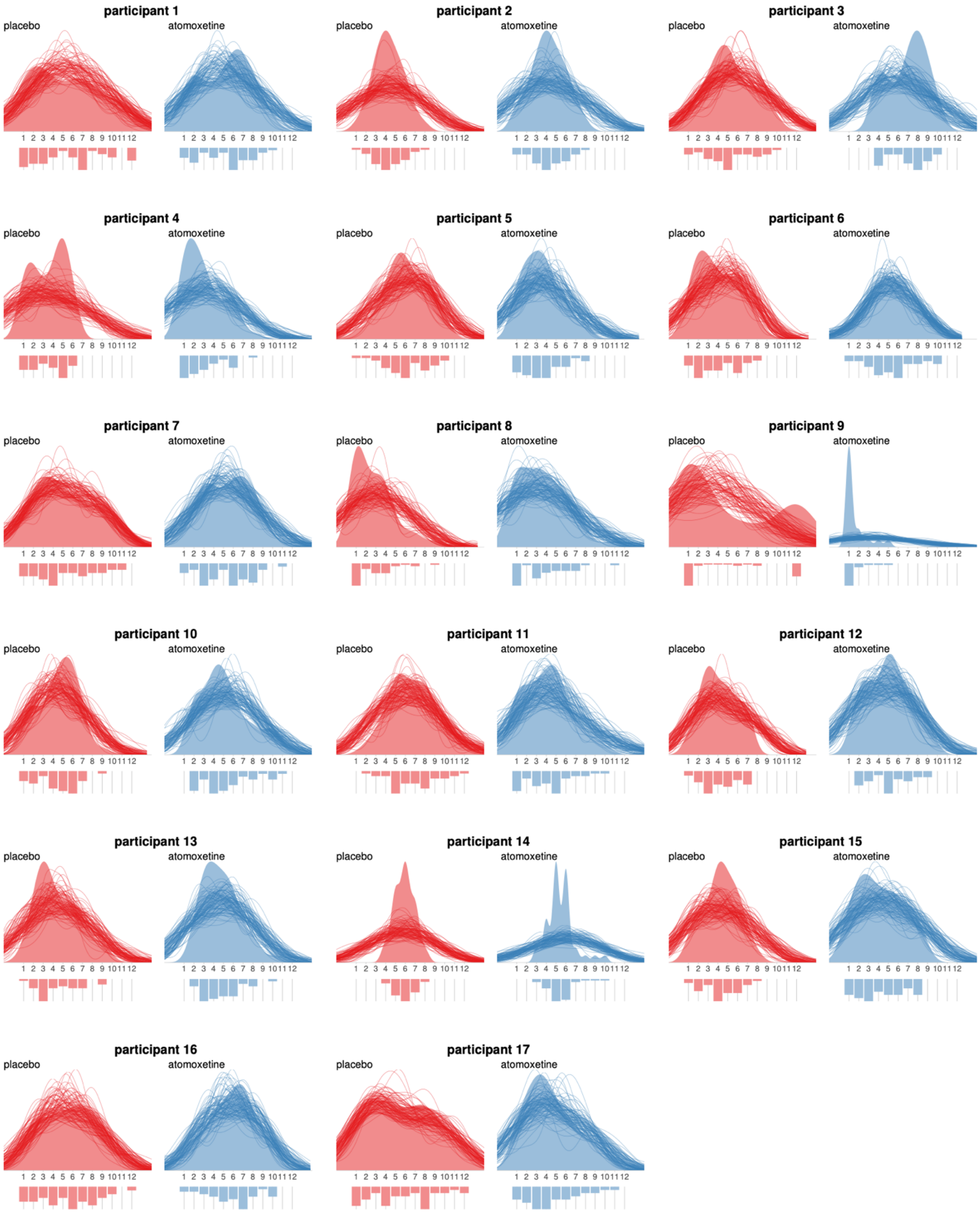

**Figure S7 | Posterior predictive check: response distributions.** Each panel compares the observed responses (light-coloured density plot and histogram) to distributions of simulated responses drawn from the model's posterior predictive distribution (dark-coloured density traces).

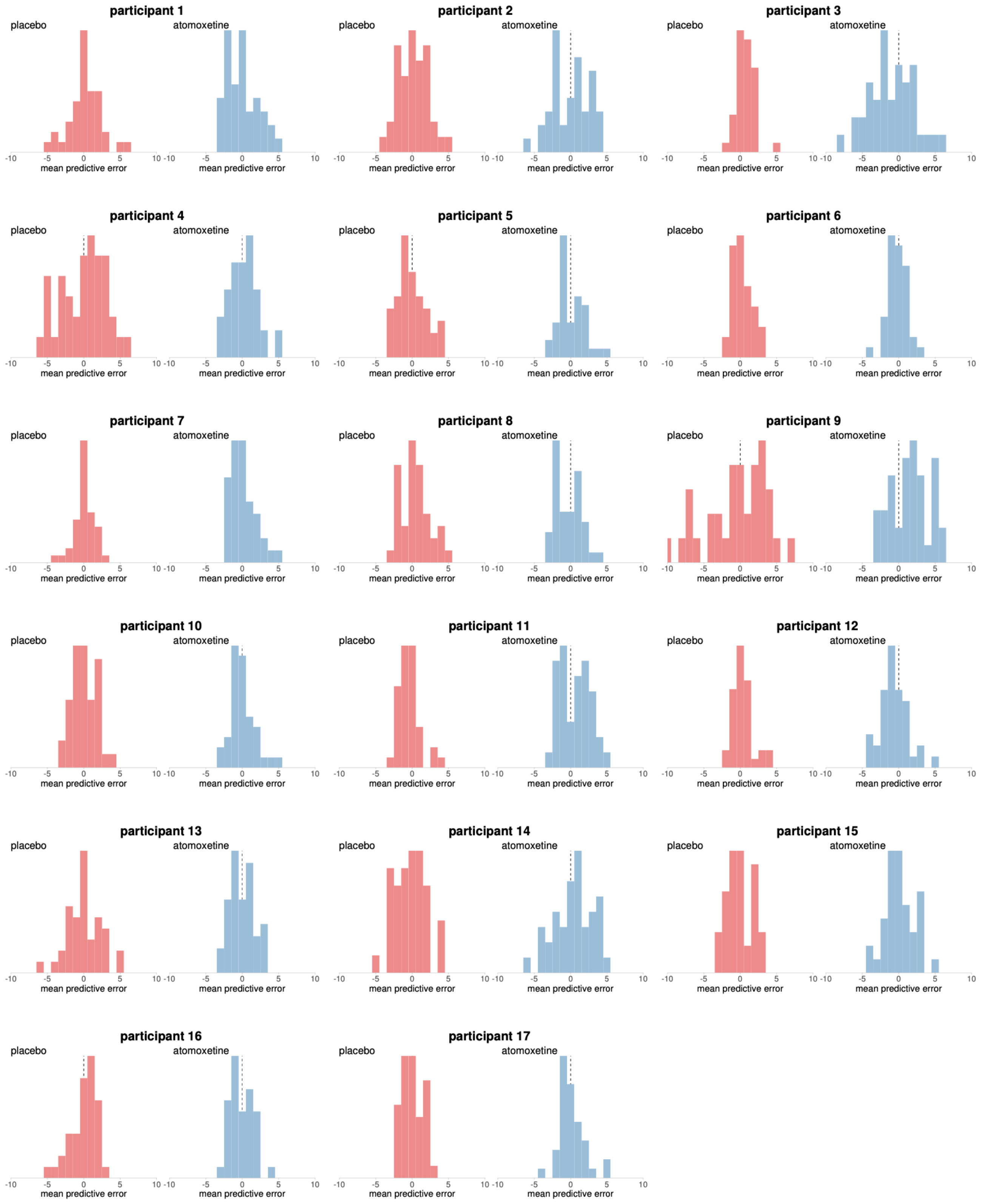

**Figure S8 | Posterior predictive check: predictive error.** Each panel illustrates the distribution of the mean predictive error of the model – that is, the observed responses minus simulated responses drawn from the model's posterior predictive distribution, averaged across Markov Chain Monte Carlo samples. These histograms can therefore be interpreted as the distributions of residuals.
